# A Protocol Describing A Scoping Review to Characterise ICU Admission Criteria

**DOI:** 10.1101/2023.10.24.23297448

**Authors:** James Soares, Chris Andersen

## Abstract

**Introduction:** Effectively identifying deteriorated patients is vital to the development and validation of automated systems designed to predict clinical deterioration. Existing outcome measures used for this purpose have significant limitations. Published criteria for admission to high acuity inpatient areas may represent markers of patient deterioration and could inform the development of alternate outcome measures.

**Objectives:** This is a protocol for a scoping review which aims to characterise published criteria for admission of adult inpatients to high acuity inpatient areas including intensive care units.

**Data sources:** Electronic databases *PubMed* and *ProQuest EBook Central* will be searched to identify papers published from 1999 to date of search. Publications of interest are those which describe prescriptive criteria for admission of adult inpatients to a clinical area with a higher level of care than a general hospital ward.

**Charting methods:** Data will be extracted from each publication using a standardised data-charting form.

**Data synthesis:** Admission criteria characteristics from included publications will be summarised and presented in text and summary table form.

## Introduction

Unrecognised clinical deterioration is a significant cause of reversible morbidity and mortality in hospitalised patients.^1–3^ Effective identification of deteriorated patients is imperative to the development of automated systems designed to predict and prevent clinical deterioration. Presently, there is little agreement in the literature on criteria to define the deteriorated ward patient^11^ and existing proxy measures of clinical deterioration – in-hospital mortality, in-hospital-cardiac-arrest (IHCA) and/or unplanned intensive care unit (ICU) transfer – have limitations which introduce subjectivity and variability to studies that use them as outcome measures.^4^ The present study aims to better characterise how deterioration has been conceptualised/defined by clinicians in practice, in order to inform the development of alternate outcome measures. To do this, we will conduct a scoping review to identify criteria that have been used to determine when hospital inpatients are unwell enough to warrant admission to a higher acuity non-ward environment such as the intensive care unit (ICU) or high depency unit (HDU).

## Objective

To describe published criteria for admission of adult inpatients to ICU or other clinical care areas with more intensive medical or nursing care than a general ward. A secondary aim will be to identify variables, among the identified criteria, that are potentially extractable from the electronic health record.

## Methods

This scoping review will conform to the requirements of the Preferred Reporting Items for Systematic reviews and Meta-Analyses extension for Scoping Reviews (PRISMA-ScR) guidelines.^12^

### Search strategy

Relevant studies will be identified by searching the electronic databases *PubMed* and *ProQuest EBook Central*. Citations of included publications will be reviewed to source additional papers.

The following search terms will be included: intensive care, intensive care unit, medical intensive care, surgical intensive care, acute care, critical care, high dependency unit, hdu, coronary care unit, admission, disposition, transfer, triage, criteria, policy, guideline, decision, selection, threshold, deteriorating, deteriorated, unstable, acute, rapid response.

Additional terms will be considered and search strategy refined after initial trials with these terms. A trained librarian from the University of New South Wales will assist with construction of the search strategy after initial trials.

### Inclusion criteria

Quantitative or qualitative studies in peer-reviewed journals, practice guidelines or book chapters which describe prescriptive criteria for admission or transfer of adult (>16 years) inpatients to ICU/HDU (including close-observation units, coronary care units and step-down units). Studies published from January 1999 to date of search will be included. No language restrictions will be applied. Google Translate will be used to translate non-English studies to English.

### Exclusion criteria

Abstract-only reports, articles including paediatric subjects (<16 years old), articles where criteria were derived a posteriori and articles focusing only on ICU exclusion criteria will be excluded.

### Selection of source and data charting

Two reviewers will independently screen titles +/- abstracts of studies identified by initial search. Disagreement regarding study eligibility will be resolved by a nominated third party. A standardised data-charting form will be used to extract the following information from each publication: type; author(s); year of publication; study objective, population and primary outcome; number of subjects; setting; method of data analysis; nature of evidence underpinning admission criteria; and characteristics of proposed admission criteria.

### Data Synthesis

We aim to generate a list of admission criteria and thresholds for ICU admission from included studies. We will chart the frequency with which each criterion is proposed as potential indicator of consensus in the literature. We will also attempt to categorise these criteria and describe themes relating to their characteristics. We will present our findings in text and summary table form.

## Data Availability

All data produced in the present work are contained in the manuscript.

